# Efficacy and safety of tirofiban following intravenous thrombolysis: A systematic review and meta-analysis of randomized controlled trials

**DOI:** 10.1101/2025.11.04.25339551

**Authors:** Sher Bano, Faisal Wali Ahmed, Atif Saeed, Ammar Nawazish, Muhammad Haris, Ayesha Mansoor, Ayesha Ghazal Jamali, Riaz Ahmed, Shaumile Hasan Khan, Numan Akram, Zainab Salahuddin, Javed Iqbal, Asraf Hussain

## Abstract

**Background:** The risk of re-occlusion and inadequate reperfusion limits the efficacy of thrombolysis within 4.5 hours for acute ischemic stroke. Tirofiban, a glycoprotein IIb/IIIa inhibitor, has been studied as an adjunctive treatment for patients with acute coronary syndrome. This meta-analysis evaluates existing evidence on the effectiveness and safety of tirofiban following thrombolysis in patients with acute ischemic stroke.

**Methods:** PubMed, Science Direct, Clinicaltrials.gov, and The Cochrane Library were searched for RCTs between 2011 and 2025. All studies included acute ischemic stroke patients, aged ≥ 18 years, NIHSS >4, who received IV thrombolysis before tirofiban administration. Endpoints included mRS score, SICH, risk of systemic bleeding, and mortality. The quality of the studies was assessed using the ROB2 tool, and risk ratios were calculated and analyzed. The protocol was registered on PROSPERO (CRD420251106033).

**Results:** Only five trials were included from the original articles retrieved from the searched databases. Two trials had a low risk of bias, one had some concerns, and two had a high risk of bias. Results showed a significant improvement in mRS score with IVT plus tirofiban (RR 1.32; 95% CI 1.07–1.62; p = 0.010), while also showing an increase in SICH with IVT plus tirofiban; however, the results were not significant (RR 1.86; 95% CI 0.18–19.24; p = 0.60). No difference in mortality was observed between the two groups (RR 1.06; 95% CI 0.56–2.04; p = 0.85).

**Conclusion:** IVT plus tirofiban significantly improves functional outcomes without increasing mortality despite the risk of SICH.

## Introduction

Acute Ischemic Stroke (AIS) is considered to be one of the leading causes of death among Americans, as approximately one in 19 people suffer from this pathology and end up losing their lives. (1). It poses a significant financial burden on the healthcare system, as well as nursing people back to life after having suffered an ischemic stroke, which requires emotional, physical, and monetary strength for the suffering families. Globally, stroke of ischemic etiology comprises approximately 87% of all stroke cases, hence, inducing a substantial burden on the healthcare facilities all around the world. (2). AIS occurs when a major vessel supplying blood to the brain develops an infarct, and the area being supplied becomes ischemic, resulting in neuronal damage and loss of body function, unless the affected area is reperfused using various techniques.

Currently, the treatment modality being widely used to treat patients reporting with neurological deficits secondary to AIS is Intravenous Thrombolysis (IVT) using reactivated Tissue Plasminogen Activator (rt-PA). Thrombolysis, achieved either by alteplase or tenecteplase, helps restore perfusion by lysing the clots and allowing for adequate re-canalization of the occluding vessels. This modality is most commonly used in large vessel occlusion and helps restore perfusion in areas that are much-needed. (3). However, there are certain areas where the efficacy of IVT is questioned. Research suggests that even after adequate re-canalization, almost 14-34% of the patients are unable to restore neurological function due to re-occlusion or formation of recurrent arterial thrombi. (4). Moreover, IVT is also associated with intracerebral hemorrhage, further worsening the functional outcomes of the concerned patients. (5).

Recently, many other pharmacological agents have been explored that inhibit re-thrombosis and increase the efficacy and safety profile of mechanical options when used in conjunction. Among these options, the use of antiplatelet agents is extremely popular these days as they inhibit the formation of microthrombi after mechanical treatment options such as IVT. Among these options, Tirofiban has been gaining considerable interest as it is a reversible platelet surface glycoprotein inhibitor that inhibits platelet aggregation, thereby reducing clot formation. (6). It has a shorter half-life and a reversible mode of action, making it suitable for acute infarcts and acute states such as AIS and other acute coronary syndromes.

Currently, many studies are being conducted to explore the role of Tirofiban in the management of Acute Ischemic Stroke, especially in cases where IVT of mechanical Thrombectomy has remained unsuccessful or partially effective. Many studies suggest that there is a noticeable improvement in neurological functional score and the general outcomes of the patients who have received Tirofiban after IVT. (7). Moreover, there was no increased risk of intracranial hemorrhage associated with Tirofiban in conjunction with IVT compared to IVT alone. However, despite raging evidence regarding the importance of Tirofiban therapy in AIS, there are some concerns regarding its safety profile and the association of this agent with intracranial bleeding a few days after the procedure. (8).

Heterogeneity in study design, patient groups, dosing regimens for Tirofiban, and definitions of outcomes makes individual results difficult to interpret. Most of the available evidence comes from single-center studies or post-hoc analyses with small patient numbers and poor quality of care. A limited consensus exists on the optimal timing, dose, and patient selection criteria; therefore, it is necessary to conduct a systematic review of the available data to inform these decisions. Consequently, it is essential to understand and interpret the safety and efficacy of this treatment modality in acute ischemic stroke patients.

Given the findings discussed above, it is a matter of hours to establish analytical evidence regarding the efficacy and superiority of Tirofiban in the prevention of secondary outcomes after an initial episode of acute ischemic stroke, to make this treatment regimen an essential part of clinical practice all around the globe, and ensure a homogeneity in guidelines addressing stroke prevention and recurrence. The primary aim of conducting this meta-analysis is to evaluate and contextualize the effectiveness of Tirofiban in combination with IVT in patients diagnosed with acute ischemic stroke, to inform the development of current clinical guidelines and provide optimal healthcare to patients presenting with this condition.

## Materials and Methods

This meta-analysis has been conducted according to the PRISMA (Preferred Reporting Items for Systematic reviews and Meta-Analyses) guidelines, and the review protocol has been registered on PROSPERO under the ID CRD420251106033 (9).

### Search Strategy

We selected three central databases to search for articles to be screened for inclusion in this review. These databases included PubMed, Science Direct, and The Cochrane Library. Specific keywords, utilizing Boolean operators "OR" and "AND", were designed to search for articles from 2011 to 2025 in an extensive manner. No other filters were applied. The keywords used were: "((((tirofiban) OR (Glycoprotein IIb/IIIa)) OR (GP IIb/IIIa)) AND (thrombolysis)) AND (Acute Ischemic Stroke)". Exact keywords were used for all the databases.

### Eligibility Criteria

A selection criterion was designed against which the articles were to be screened. The key inclusion criteria included patients of acute ischemic stroke having an age above 18 years who underwent thrombolysis within 4.5 hours and were given tirofiban within 24 hours of thrombolysis. An NIHSS score of greater than 4 at baseline was required for studies to be included. Only randomized controlled trials published in English were included. Studies were included if they reported on any of the following outcomes: modified Rankin scale scores, symptomatic intracranial hemorrhage, mortality, and systemic bleeding.

Studies were excluded for having an age below 18 years, no evidence of thrombolysis before tirofiban, patients undergoing endovascular thrombectomy, NIHSS score <3, patients having hemorrhagic stroke, or tirofiban after 24 hours of IVT. Study designs other than RCTs were excluded, and studies published in any language other than English were excluded as well.

Studies without any of the outcomes mentioned above were also excluded.

### Screening, Selection, and Data Extraction

Initially, all the retrieved articles were searched for duplicates and removed. The remaining articles underwent title and abstract screening, after which the selected articles were searched for full texts. The full texts were then screened against the selection criteria. Two reviewers carried out this process of screening and selection. If any dispute arose regarding the selection of an article, a third reviewer made the final decision regarding its selection. Articles that fulfilled the selection criteria were then included and underwent data extraction and synthesis.

Data extraction was performed independently by two reviewers. The data were extracted into an Excel sheet and included study ID, location, sample size, clinical trial number, mean age, number of males and females, intervention details, comparator details, outcomes, and characteristics of participants.

### Quality Assessment and Data Synthesis

The ROB2 tool, developed by The Cochrane Library, was used to assess the quality of the included trials. (10). This tool consists of 5 domains, with biases arising from randomization, selection bias, bias due to missing outcome data, bias due to deviating from the intervention, and prejudice in outcome measurement. The studies were considered low risk if all domains had a low risk of bias, while they were regarded as having some concerns if any domain had a high risk. In contrast, one domain raised concerns, and studies were considered high risk if they either had one domain with high risk or at least two domains with some reservations.

RevMan 5.4 was used to analyze the data collected for the outcomes. Risk ratios were calculated for dichotomous variables; a random—effects Mantel-Haenszel model was used. I2 was used as a measure of statistical heterogeneity, with I2<25 considered as low, 26-50 as mild, 51-75 as moderate, and >75 as high. Forest plots were used to represent results for the specified outcomes. A funnel plot for publication bias was not used due to the low number of studies included. A p-value below 0.05 was considered significant. A leave-one-out sensitivity analysis was conducted to identify the sources of heterogeneity and to assess the robustness of our findings. Subgroup analysis couldn’t be undertaken due to the limited data available.

## Results

Initially, 2055 articles were retrieved using specified keywords from the included databases. Firstly, duplicates were removed, and the remaining 1999 articles underwent title and abstract screening, leaving 48 articles behind. The full text of 9 articles was not found, and the selection criteria were applied to the remaining 39 articles. Only five articles fulfilled the selection criteria and were included in our final analysis. (11, 12, 13, 14, 15). The PRISMA flowchart for screening and selection, along with reasons for excluding articles, is shown below in Fig 1.

**Figure 1.**
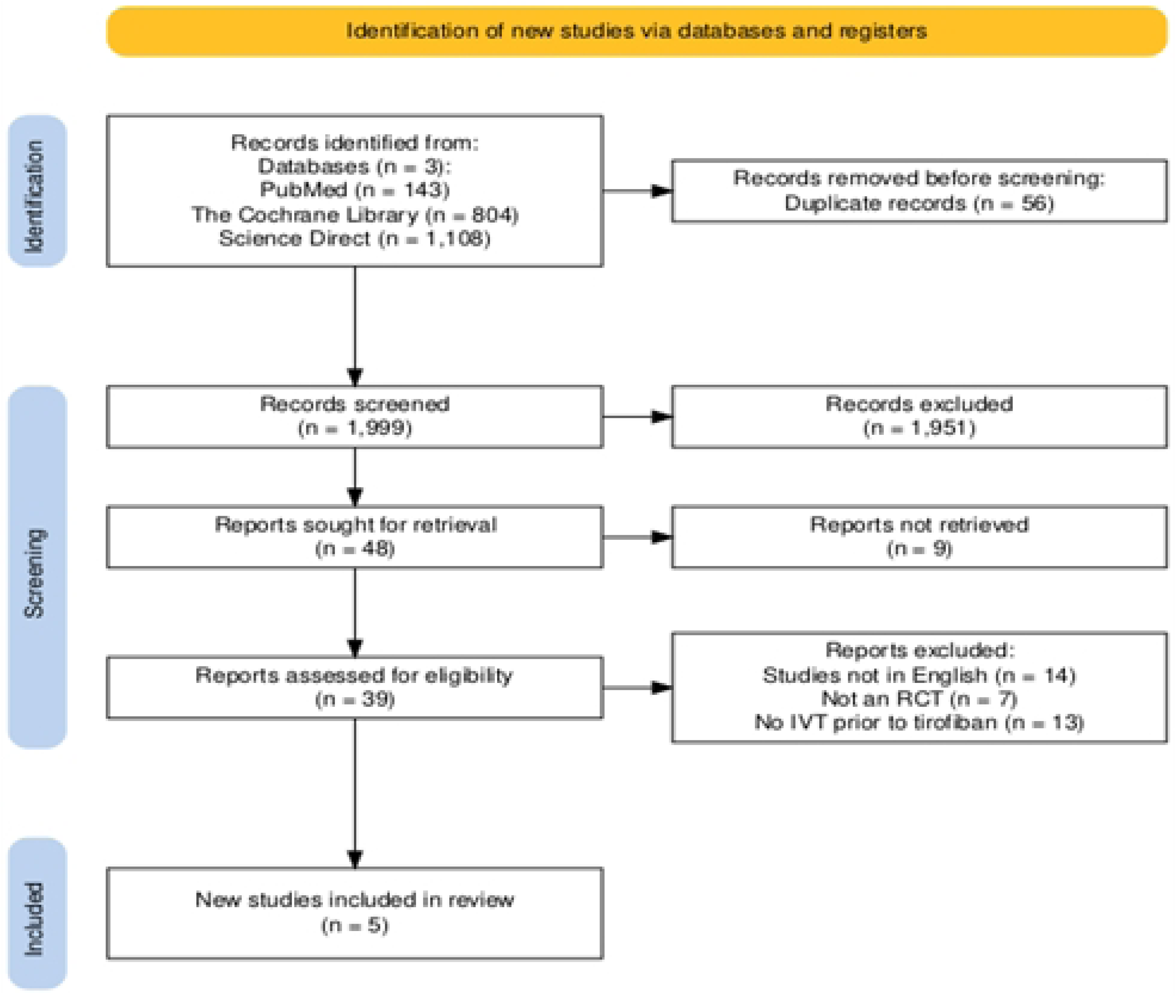
PRISMA flow diagram of study selection. Flow diagram showing the process of database searching, screening, eligibility assessment, and inclusion of randomized controlled trials.

### Characteristics of Studies

This review included five trials with a total sample of 1224 participants, out of which 789 (64.5%) were males and 435 (35.5%) were females. The majority of the population was hypertensive (77.7%), while 309 (25.2%) were diabetics and 270 (22%) had a previous history of stroke or TIA. NIHSS score was above 4 in all the studies, and alteplase was mainly given for thrombolysis within 4.5 hours, except for Tao C et al., 2025, where tenecteplase and alteplase were used (14). 721 (58.9%) participants were randomized to the tirofiban plus IVT group, while 503 (41.1%) participants were included in the IVT alone group. The characteristics of the included studies are summarized in Table 1.

**Table 1:** Characteristics of the included randomized controlled trials.

|  | Included Studies |  |  |  |  |
| --- | --- | --- | --- | --- | --- |
|  | Tao C et al., 2025 | Zhang Y et al., 2022 | Liang Z et al., 2022 | Liu J et al., 2019 | Li W et al., 2016 |
| **Clinical Trial Number** | NCT06134622 | ChiCTR2200058513 | ChiCTR1800014666 | - | ChiCTR-TRC-14004630 |
| **Sample Size** | 832 | 73 | 60 | 177 | 82 |
| **Age** | Tirofiban: 69 (59-76)<br>Placebo: 69 (59-76) | Tirofiban: 69.24 ± 14.88<br>Placebo: 68.21 ± 12.86 | Tirofiban: 65.33 ± 8.18<br>Placebo: 66.53 ± 10.27 | Tirofiban: (a) 66.26 ± 6.01<br>(b) 68.05 ± 8.25<br>(c) 67.90 ± 6.83<br>Placebo: 67.71 ± 6.72 | Tirofiban: 66 (29-86)<br>Placebo: 68 (37-86) |
| **Male: Female** | 531: 301 | 39: 34 | 38: 22 | 132: 45 | 49: 33 |
| **Comorbidities** |  |  |  |  |  |
| **Hypertension** | 678 | 57 | 39 | 129 | 48 |
| **Diabetes Mellitus** | 191 | 14 | 12 | 70 | 22 |
| **Stroke or TIA** | 230 | 11 | - | 22 | 7 |
| **Hyperlipidemia** | 109 | 17 | - | 70 | - |
| **IHD** | 90 | 15 | - | 17 | 11 |
| **NIHSS (tirofiban)** | 6 (5-9) | 8.90 ± 2.75 | 7.63 ± 3.54 | (a) 10.11 ± 5.04<br>(b) 9.25 ± 4.52<br>(c) 9.10 ± 4.38 | 8 (4–18) |
| **Time to tirofiban** | 44 mins<br>(27-55) | Within 24 hours | Within 24 hours | (a) Within 2 hours of IVT<br>(b) Within 2-12 hours of IVT (c)<br>Within<br>12-24<br>hours of<br>IVT | Within 24 hours |
| **Intervention**<br>**(Tirofiban)** | 0.4 µg/kg<br>OF<br>tirofiban for<br>30 minutes,<br>then a<br>maintenanc<br>e dose of<br>0.1<br>µg/kg/min<br>for 23.5<br>hours | 0.4 µg/kg OF<br>tirofiban for 30<br>minutes, then a<br>maintenance<br>dose of 0.1<br>µg/kg/min for at<br>least 24 hours | 0.4 µg/kg OF<br>tirofiban for 30<br>minutes, then a<br>maintenance<br>dose of 0.1<br>µg/kg/min for at<br>least 24 hours | 5 µg/kg<br>of<br>tirofiban<br>bolus,<br>then<br>maintena<br>nce dose<br>of 0.1<br>µg/kg/mi<br>n for 24 h | 0.4 µg/kg<br>OF<br>tirofiban<br>for 30<br>minutes,<br>then a<br>maintena<br>nce dose<br>of 0.1<br>µg/kg/mi<br>n for at<br>least 24<br>hours |
| **Outcomes** | mRS<br>scores, ED-QOL,<br>NIHSS,<br>SICH, | mRS scores,<br>SICH, mortality,<br>systemic<br>bleeding | mRS scores,<br>SICH, systemic<br>bleeding | mRS<br>scores,<br>SICH | mRS<br>scores,<br>SICH,<br>mortality,<br>systemic |
|  | mortality |  |  |  | bleeding |

### Risk of Bias

Of the five included trials, only two had an overall low risk of bias. The study by Liang Z et al. (2022) raised concerns about the randomization process, which in turn raised concerns about the overall validity of the research. (11). While the studies by Liu J, et al. 2019 and Zhang Y, et al. 2022 had an overall high risk of bias, with some concerns in the randomization process and high risk in deviations from interventions for Liu J, et al. 2019, and some problems in domains 2,3,4 for Zhang Y, et al. 2022 (13, 15). The detailed risk of bias assessment is illustrated in Fig 2.

**Fig 2.**
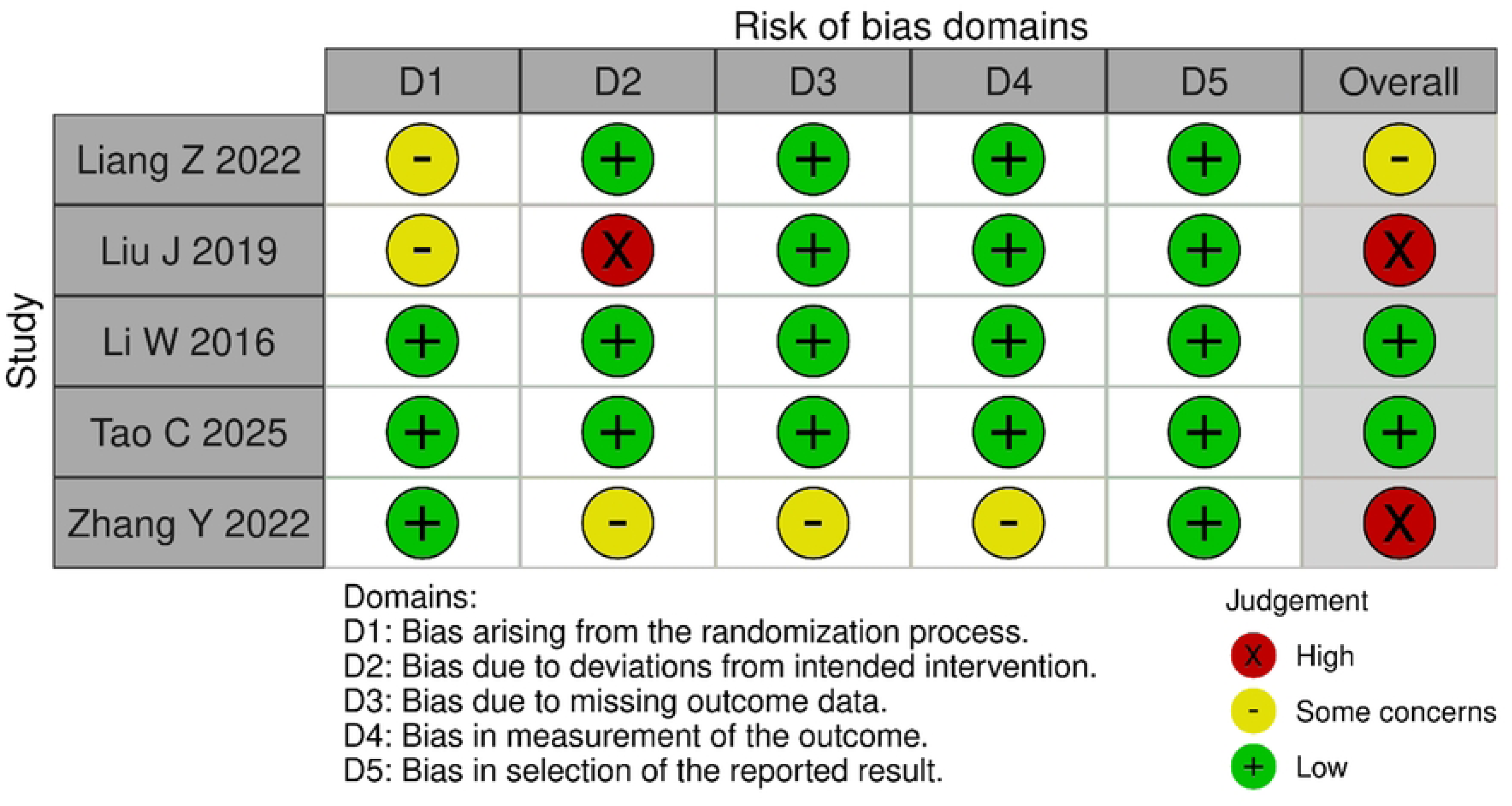
Risk of bias assessment. Risk of bias summary for included trials using the Cochrane RoB 2 tool.

### mRS Scores

All the trials included in this review reported on functional outcome using mRS scores. Analysis of the data on the mRS score of 0-2 showed that participants in the IVT-only group had a significantly poorer functional outcome compared to the experimental group receiving tirofiban after IVT (RR 1.32; 95% CI 1.07–1.62; p = 0.010). Despite the significant results, moderate statistical heterogeneity was observed, with an I2 value of 70%. The forest plot of mRS scores is presented in Fig 3.

**Fig 3.**
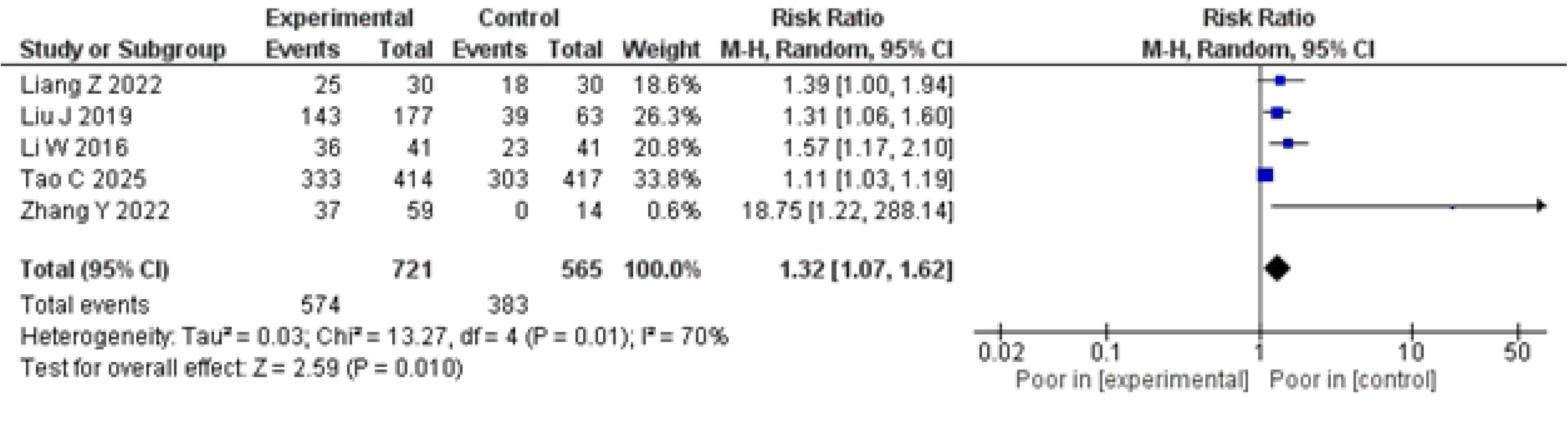
Forest plot of functional outcome (mRS 0–2). Comparison of favorable outcomes (mRS 0–2) between IV thrombolysis alone and IV thrombolysis plus tirofiban.

### SICH and Systemic Bleeding

Only four trials reported on the outcomes of SICH and systemic bleeding, with Liang Z, et al. (2020) having zero events of SICH and systemic bleeding in both the experimental and control groups. For SICH, the risk was increased in the tirofiban plus IVT group compared to the IVT group (RR 1.86; 95% CI 0.18–19.24; p = 0.60), but the result was not statistically significant. However, mild statistical heterogeneity was observed, with an I2 value of 50%. The forest plot for SICH is shown in Fig 4.

**Fig 4.**
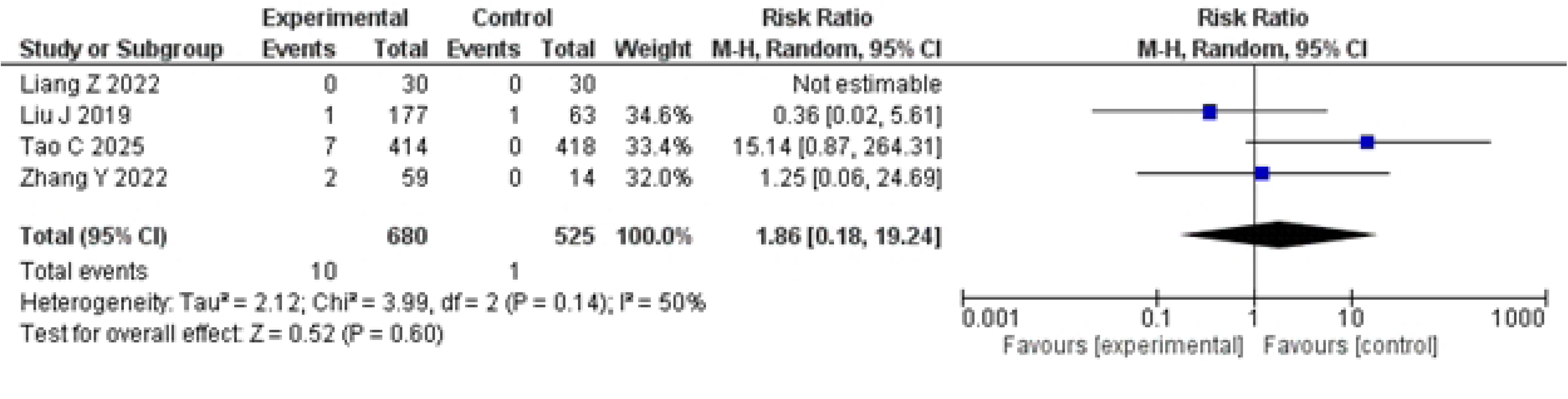
Forest plot of symptomatic intracranial hemorrhage (SICH). Risk ratio for SICH in IV thrombolysis alone versus IV thrombolysis plus tirofiban groups.

For systemic bleeding, the risk was reduced in the tirofiban plus IVT group compared to the IVT group, but the result for SICH was not significant (RR 0.84; 95% CI 0.38–1.86; p = 0.67), with no heterogeneity. The forest plot for systemic bleeding is shown in Fig 5.

**Fig 5.**
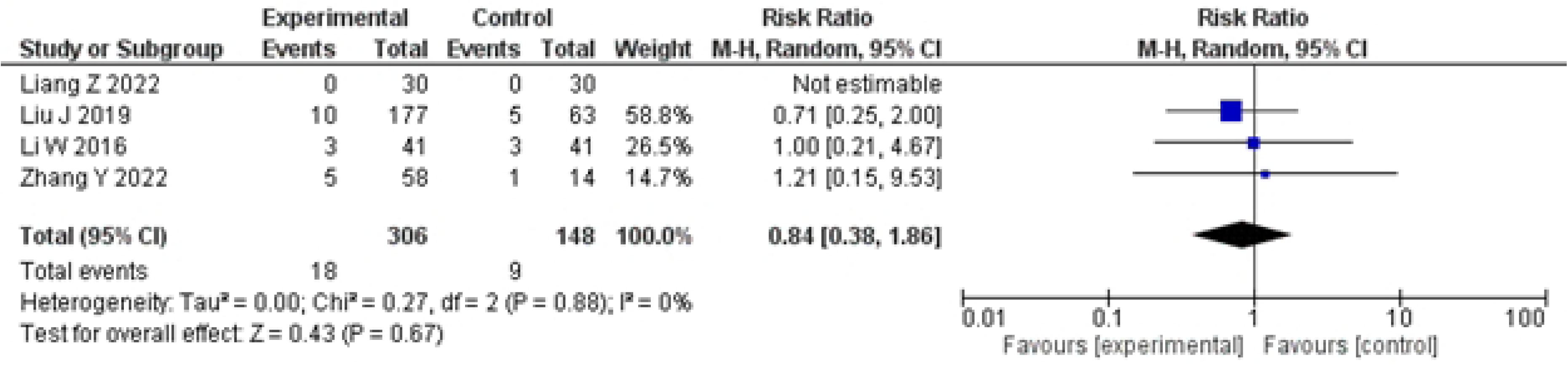
Forest plot of systemic bleeding. Comparison of systemic bleeding risk between IV thrombolysis alone and IV thrombolysis plus tirofiban.

### Mortality

All five trials reported mortality outcomes; however, no mortality was observed in either the experimental or the control group in three of the studies. Among the two studies that reported mortalities, no significant difference was observed between the two groups (RR 1.06; 95% CI 0.56–2.04; p = 0.85), with no heterogeneity. The forest plot for mortality is shown in Fig 6.

**Fig 6.**
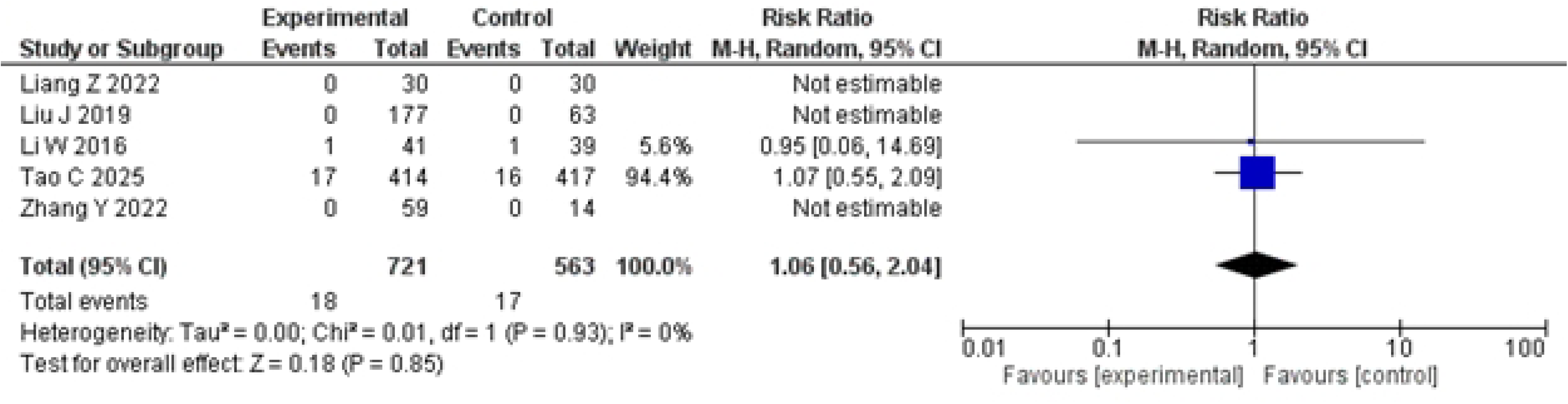
Forest plot of mortality. Risk ratio for mortality between the IV thrombolysis alone and the IV thrombolysis plus tirofiban groups.

### Sensitivity Analysis

No significant results were observed after performing sensitivity analysis; however, for mRS scores, the sensitivity analysis showed that removal of Tao C, et al. 2025 which had the most significant number of participants reduced heterogeneity from moderate to mild while the results remained significant in favor of tirofiban plus IVT group (RR 1.43; 95% CI 1.13–1.82; p = 0.003; I2=47%).

For SICH, the results favored IVT plus tirofiban compared to IVT alone after excluding Tao C, et al. (2025), but remained insignificant with no heterogeneity (RR 0.63; 95% CI 0.08–4.81; p = 0.54; I2 = 0%). For systemic bleeding and mortality, the results didn’t change significantly upon performing sensitivity analysis.

## Discussion

Our study evaluates the efficacy and safety of tirofiban administration after IV thrombolysis in acute ischemic stroke by investigating and analyzing functional outcomes using mRS scores, as well as safety outcomes, including SICH, systemic bleeding, and mortality rate. The experimental groups that received tirofiban after IV thrombolysis had a better functional outcome than those that received only IV thrombolysis, with these results being statistically significant. Receiving tirofiban after IV thrombolysis increased the risk of SICH; however, the sensitivity analysis showed the opposite result, with a decreased risk in the tirofiban group, yet the outcome remained insignificant. Patients who did not receive tirofiban after IV thrombolysis had a higher chance of systemic bleeding. No significant difference was observed between the two groups in mortality outcomes.

Intravenous thrombolysis is widely accepted as a treatment modality for acute ischemic stroke, but limited access in different countries calls for looking into other effective treatments. Tirofiban reduces platelet aggregation by inhibiting the GP IIb/IIIa receptor, thereby preventing fibrinogen from binding to it. This reduces early platelet-driven re-occlusion of the vessel post-thrombolysis (12, 16). This phenomenon may be the reason for the better functional outcome observed in our analysis, among the participants who received tirofiban after IV thrombolysis. A similar result was also observed in another meta-analysis by Liu et al., where the tirofiban group with IVT had better mRS scores. (7). These results are in line with previous studies on a similar topic, which suggest that tirofiban administration is associated with improvements in clinical outcomes and overall neurological function assessment scores in patients with acute ischemic stroke. (17).

However, the presence of moderate heterogeneity (I² = 70%) indicates variability across studies. Sensitivity analysis revealed that the removal of the survey by Tao et al. (2025) reduced heterogeneity to a mild level (I² = 47%), while maintaining statistical significance (RR 1.43; 95% CI 1.13–1.82; p = 0.003). This reinforces the robustness of the primary outcome and suggests that the overall positive effect of Tirofiban is not driven solely by a single large trial. Nevertheless, the variation across studies in terms of patient selection, Tirofiban dosage, timing of administration, and stroke severity must be considered when interpreting these findings, as these factors can significantly impact the results. (18).

Similarly, the risk of SICH, when evaluated with Tirofiban administration, also showed heterogeneity, as the results were clinically not significant in the group receiving Tirofiban and IVT in combination. This is in accordance with research conducted before that suggested that Tirofiban tended to increase bleeding events when given in combination with IVT. However, after the removal of Tao et al.’s 2025 study, the risk ratio shifted in favor of tirofiban, thus indicating and reinforcing the fact that tirofiban can prove beneficial in patients under controlled conditions. (19).

Our meta-analysis revealed no significant difference in death rates between tirofiban and intravenous thrombolysis versus IVT alone. Tirofiban caused slightly less systemic bleeding than thrombolytics, most likely because thrombolytics destroy clots throughout the body, whereas tirofiban inhibits platelet aggregation, increasing bleeding risk in vulnerable areas. (20).

However, extracranial bleeds are frequently less dangerous than intracranial hemorrhage, which is the leading cause of death following thrombolysis. (21). Without a considerable reduction in ICH, the survival benefit of fewer systemic bleedings is limited. (22).

## Limitations

Certain limitations need to be acknowledged. Firstly, our study analyzed a limited number of studies, which may potentially disrupt the results and exclude events caused by rare occurrences. Only two studies were classified as having a low overall risk of bias, with the rest having issues related to randomization, deviations from intended interventions, and the reporting of outcomes. This suggests that high-quality, appropriately powered randomized controlled trials are necessary to determine the role of tirofiban in AIS management conclusively. It is notable, however, that the observed effect persisted even when studies of varying methodological quality were included, suggesting a possibly durable effect. Similarly, there was a difference in the number of participants based on gender. As some drugs may function better in males, the male prevalence could have altered the results. The failure to perform subgroup analysis due to limited data available is an additional limitation.

## Conclusion

In conclusion, this meta-analysis demonstrates that tirofiban administration after IVT procedure in ischemic stroke is associated with improved functional outcomes without a significant increase in SICH or mortality. These findings suggest that tirofiban is a good alternative in selected AIS patients. However, further research is needed to quantify its specific dosage and to incorporate it into universal guidelines for stroke treatment. This research paves the way for future researchers to explore the optimal dosage of Tirofiban needed for adequate Thrombolysis and the prevention of re-thrombosis in patients post-IVT. Similarly, there is a need of the hour to classify the efficacy of this treatment modality based on stroke subtype and the etiology behind its occurrence.

## Acknowledgments

## Author Contributions

Conceptualization: Sher Bano.

Methodology: Sher Bano, Faisal Wali Ahmed, Atif Saeed. Investigation: Ammar Nawazish, Muhammad Haris.

Formal analysis: Ayesha Mansoor, Ayesha Ghazal Jamali. Writing – original draft: Riaz Ahmed, Shaumile Hasan Khan.

Writing – review & editing: Numan Akram, Zainab Salahuddin, Javed Iqbal, Asraf Hussain. Data curation: Ammar Nawazish, Muhammad Haris.

Supervision: Asraf Hussain.

## Data Availability

All relevant data are within the manuscript and its Supporting Information files.

